# Modelling the Interplay between Responsive Individual Vaccination Decisions and the Spread of SARS-CoV-2

**DOI:** 10.1101/2023.08.24.23294588

**Authors:** Karina Wallrafen-Sam, Maria Garcia Quesada, Benjamin A. Lopman, Samuel M. Jenness

**Affiliations:** Department of Epidemiology, Rollins School of Public Health, Emory University, Atlanta, Georgia, USA

**Author notes:** **CORRESPONDENCE** Karina Wallrafen-Sam, Emory University, 1518 Clifton Road, Atlanta, GA, 30322, USA. **FUNDING**. **DISCLAIMER**. The findings and conclusions in this report are those of the authors and do not necessarily represent the official position of the funding agencies. **CONTRIBUTIONS**. KWS conceived of, designed, and conducted the analysis and wrote the manuscript. SMJ provided direction to the development of the study design, supervised the analysis, and provided critical input to the manuscript. BAL and MGQ provided input on the study design and critically reviewed and edited the manuscript. **CONFLICTS OF INTEREST**.

## Abstract

The uptake of COVID-19 vaccines remains low despite their high effectiveness. Epidemic models that represent decision-making psychology can provide insight into the potential impact of vaccine promotion interventions in the context of the COVID-19 pandemic. We coupled a network-based mathematical model of SARS-CoV-2 transmission in Georgia, USA with a social-psychological vaccination decision-making model in which vaccine side effects, post-vaccination infections, and other unidentified community-level factors could “nudge” individuals towards vaccine resistance while hospitalization spikes could nudge them towards willingness.

Combining an increased probability of hospitalization-prompted resistant-to-willing switches with a decreased probability of willing-to-resistant switches prompted by unidentified community-level factors increased vaccine uptake and decreased SARS-CoV-2 incidence by as much as 30.7% and 24.0%, respectively. The latter probability had a greater impact than the former. This illustrates the disease prevention potential of vaccine promotion interventions that address community-level factors influencing decision-making and anticipate the case curve instead of reacting to it.

---

The most impactful intervention to date against the COVID-19 global pandemic has been the development and distribution of highly effective vaccines.^1,2^ The COVID-19 vaccine rollout in the United States began in late 2020 with a two-dose primary series, before waning immunity and decreased effectiveness against novel SARS-CoV-2 strains prompted the development of four sequential booster vaccinations.^3^ Although these vaccines have been widely available in the United States since 2021, only about 70% of the eligible U.S. population had completed the primary vaccine series as of mid-2023.^4^ Primary series coverage was even lower in the state of Georgia, at around 58%.^4^ Vaccine uptake slowed in the later months of 2021, has been consistently lower in younger age groups, and tapered off markedly for subsequent doses.^4^ This is partially due to vaccine hesitancy – a complex phenomenon related to concerns about the safety, efficacy, and necessity of these vaccines.^5,6^

Encouraging higher vaccine uptake by addressing vaccine hesitancy is crucial, but the effects of vaccine promotion interventions can be difficult to predict since decisions on whether and when to receive a vaccine dose are influenced by a multitude of factors – including fear of illness,^7^ altruism,^7,8^ social conformity,^9,10^ and information spread via social contacts, news outlets, or social media.^11,12^ Mathematical models can compare counterfactual scenarios and represent complex individual- and community-level behaviours, providing insight into the optimal formulation and timing of such interventions. While several models of vaccination decision-making exist, most of them consider one-time decisions; are focused on theory rather than real-world scenarios; and, most crucially, are rooted in game theory, meaning they rely on the assumption of rational actors.^13–16^ In contrast, social psychological research suggests that individuals typically rely on heuristics rather than rational cost-benefit analyses when making complex decisions.^17^ For example, decisions generally obey the law of inertia: they tend to remain stable over time but are sensitive to small “nudges” from unfavourable outcomes.^18^ To provide valid insights into vaccination interventions, mathematical models are needed that account for such heuristics and capture feedback effects between behaviour and epidemic outcomes.

The role of heuristics and inertia in decision-making has been studied in the context of annual influenza vaccination but is not yet well understood in the context of the ongoing COVID-19 vaccine rollout. Papst et al developed a seasonal influenza model in which prior infections and vaccine side effects could “nudge” persons to change their future vaccination decisions, creating a feedback loop between behaviour and disease spread.^19^ This model, while theoretical in its focus, was consistent with the empirical findings of a longitudinal cohort study on trends in influenza vaccination: the cohort’s vaccination behaviour was generally stable over time, but flu infections could influence subsequent vaccination decisions and those who did switch between approaches tended to persist in their new behaviour, in a practical illustration of the law of inertia.^20^ Adapting the modelling framework developed by Papst et al for a mathematical model of SARS-CoV-2 could provide insight into how vaccine promotion interventions should be formulated and targeted specifically to boost the stagnant levels of COVID-19 vaccine coverage.

In this study, we utilized a model of SARS-CoV-2 dynamics in the state of Georgia – a high-burden population with significant gaps in vaccine coverage – and coupled it with a social-psychological decision-making model in which vaccine side effects, breakthrough infections, and the overall state of the outbreak could “nudge” individuals to change their vaccination behaviour. Our goal was to use this novel combination of methods to explore how changes to the probabilities of individuals adapting their behaviour after experiencing one of these “nudges” might impact disease incidence.

## RESULTS

### Model Overview

Our network-based mathematical model was parameterized and calibrated to represent the COVID-19 epidemic in the state of Georgia, USA from January 2021 to August 2022. **Figure 1** shows the calibrated model’s simulated monthly infection, hospitalization, and death rates against empirical trends in Georgia, with hospitalizations peaking in August 2021 (Delta wave) and infections peaking in January 2022 (Omicron wave). There were 131.1 infections (50% SI: 129.9, 132.3), 82.0 symptomatic cases (50% SI: 81.2, 82.7), and 0.36 deaths (50% SI: 0.35, 0.38) per 100’000 person days in this model. **Figure 2** displays the model’s simulated vaccine coverage by age group and dose against the corresponding empirical coverage.

**Figure 1.**
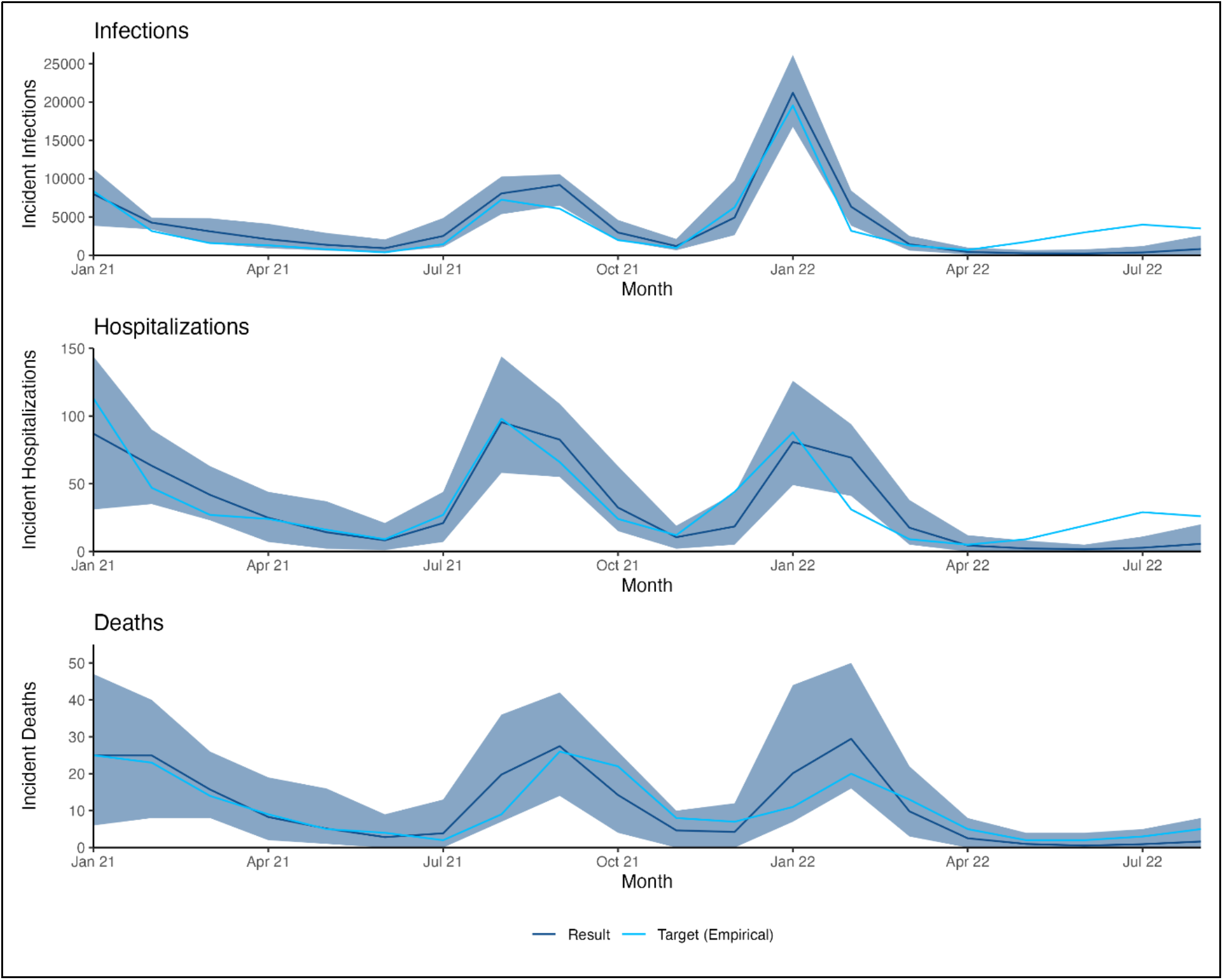
Model calibration results for cases, hospitalizations, and deaths. The reference model was calibrated to (1) an estimate of the total number of incident SARS-CoV-2 infections, (2) the reported number of confirmed COVID-19-related hospital admissions, and (3) the reported number of confirmed COVID-19-related deaths, all per 100’000 persons in Georgia per month.

**Figure 2.**
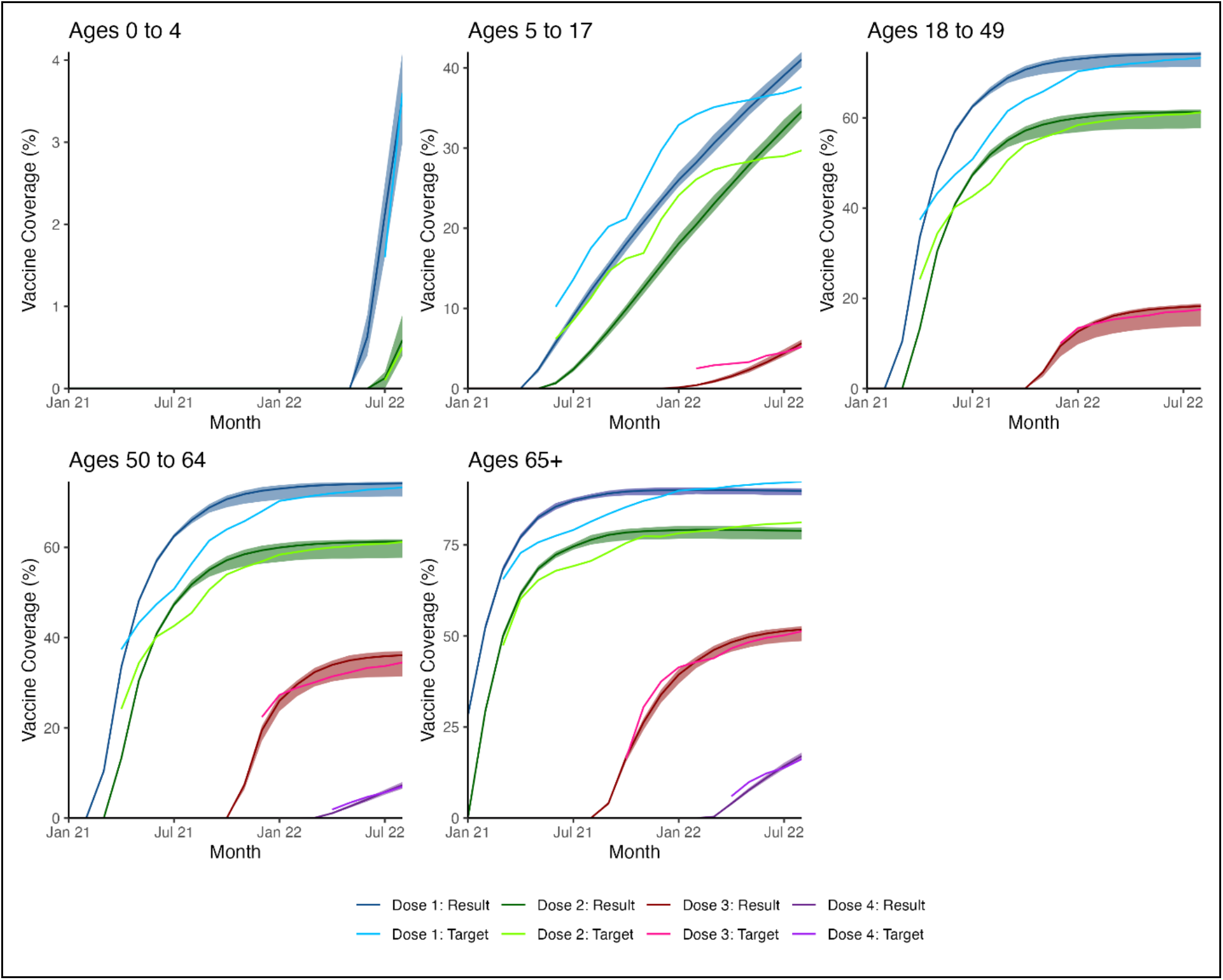
Model calibration results for vaccine coverage. The reference model was also calibrated to the reported vaccine coverage levels in Georgia by age group, dose, and month.

We compared this calibrated reference model to counterfactual scenarios in which we decreased the probability that a breakthrough infection would prompt a willing-to-resistant vaccine attitude switch (hereafter referred to as the Breakthrough Nudge Probability or Breakthrough NP); increased the probability that a spike in hospitalizations would prompt a resistant-to-willing switch (hereafter referred to as the Hospitalization Nudge Probability or Hospitalization NP); and/or decreased the probability of a vaccine willing-to-resistant switch related to something other than side effects or breakthrough infections (hereafter referred to as the Miscellaneous Nudge Probability or Miscellaneous NP). A detailed description of the model, its parameters, and the scenario specifications is provided in the Methods and in **Table 1**.

**Table 1.** Selected model parameters.

| Parameter | Value | Source |
| --- | --- | --- |
| **Population & Household Characteristics** |  |  |
| *Population proportion by age group\** | 0.233, 0.620, 0.147 | [32] |
| *Avg. household size* | 2.7 | [36] |
| *Proportion of households with members in given age group\** | 0.292, 0.791, 0.314 | [35] |
| *Proportion of children living with adult under 65* | 0.979 | [37] |
| **Community Contact Patterns** |  |  |
| *Overall daily mean degree* | 13.8 | [33] |
| *Daily mean degree for persons aged 65+* | 5.7 | [33] |
| *Associative mixing proportion by age group\** | 0.69, 0.81, 0.21 | [34] |
| **Transmission & Natural History** |  |  |
| *Reference per-act transmission probability* | 0.050 | Calibrated |
| *Relative transmission risk if asymptomatic* | 0.5 | [45] |
| *Contacts per household pairing per day* | 3 | Calculated from [46] |
| *Proportion symptomatic by age group^* | 0.573, 0.642, 0.760, 0.800, 0.813, 0.814, 0.769, 0.723, 0.666 | [47] |
| *Proportion hospitalized by age group^* | 0.006, 0.006, 0.008, 0.015, 0.021, 0.027, 0.036, 0.046, 0.054 | Calibrated; ratios from [47] |
| *Avg. duration of latent period, in days* | 5.5 | [48] |
| *Avg. duration of pre-clinical infection, in days* | 1.5 | [45] |
| *Avg. duration of clinical infection, in days* | 3.5 | [45] |
| *Avg. duration of hospitalization, in days* | 10.0 | [49] |
| *Avg. duration of asymptomatic infection, in days* | 5.0 | [45] |
| *Avg. duration of natural immunity, in days* | 300 | [42] |
| *General annual mortality rate, in deaths per 100'000 persons per year†* | 608, 30, 13, 22, 63, 116, 143, 187, 228, 300, 416, 600, 945, 1'453, 1'952, 2'817, 4'369, 7'159, 15'626 | [50] |
| *COVID-related mortality multiplier* | 1800 | Calibrated |
| **Vaccination** |  |  |
| *Proportion initially vaccine willing‡* | N/A, N/A, 0.70, 0.75, 0.91 | [38] and [39] |
| *Hospitalization nudge probability* | 0.102 | Calculated from [9] |
| *Side effect nudge probability* | 0.073 | [40] |
| *Breakthrough infection nudge probability* | 0.125 | [41] |
| *Miscellaneous nudge probability (post Dose 1)‡* | N/A, N/A, .18, .18, .12 | Calibrated |
| *Miscellaneous nudge probability (post Dose 2)‡* | N/A, N/A, .75, .43, .35 | Calibrated |
| *Miscellaneous nudge probability (post Dose 3)<sup>‡</sup>* | N/A, N/A, N/A, 0.50, 0.42 | Calibrated |
| *Hospitalization threshold in cases per 100'000* | 36 | [51] |
| *Time step for start of dose 1 rollout<sup>‡, #</sup>* | 534, 132, 84, 74, 11 | [52], [53], [54], [55], [56] |
| *Time step for start of dose 3 rollout<sup>‡, #</sup>* | N/A, 368, 323, 323, 265 | [57], [58], [59] |
| *Time step for start of dose 4 rollout<sup>‡, #</sup>* | N/A, N/A, N/A, 453, 453 | [60] |
| *Dose 1 vaccination rate, per day<sup>‡</sup>* | 0.0005, 0.0012, 0.0160, 0.0160, 0.0180 | Calibrated |
| *Dose 2 vaccination rate, per day<sup>‡</sup>* | 0.010, 0.015, 0.300, 0.300, 0.450 | Calibrated |
| *Dose 3 vaccination rate, per day<sup>‡</sup>* | N/A, 0.003, 0.045, 0.038, 0.015 | Calibrated |
| *Dose 4 vaccination rate, per day<sup>‡</sup>* | N/A, N/A, N/A, 0.005, 0.008 | Calibrated |
| *Relative risk of infection, by dose* | 0.324, 0.112, 0.120, 0.120 | [61], [62] |
| *Relative risk of symptoms, by dose* | 0.40, 0.09, 0.09, 0.09 | [63], [62] |
| *Relative risk of hospitalization, by dose* | 0.30, 0.02, 0.07, 0.07 | [63], [62] |
| *Per-dose probability of side effects* | 0.18 | [40] |
| *Half-life of vaccine immunity, in days* | 80 | [42] |

\* Values displayed for the following age groups: <18, 18-64, and 65+ yrs.
^ Values displayed for the following age groups: <10, 10-19, 20-29, 30-39, 40-49, 50-59, 60-69, 70-79, and 80+ yrs.
† Values displayed for the following age groups: <1, 1-4, 5-9, 10-14, 15-19, 20-24, 25-29, 30-34, 35-39, 40-44, 45-49, 50-54, 55-59, 60-64, 65-69, 70-74, 75-79, 80-84, and 85+ yrs.
‡ Values displayed for the following age groups: <5, 5-17, 18-49, 50-64, and 65+ yrs.

### Intervention Impacts

Doubling the Hospitalization NP generally led to a slight increase in the coverage of all vaccine doses; the absolute change ranged from about 1% to 5% (for example, for 18- to 49-year-olds, coverage changed from 74.2%, 61.4%, and 18.3% for the first, second, and third doses to 77.0%, 64.9%, and 22.8%). Reducing the Breakthrough NP to 0 had no discernible impact. In contrast, reducing the Miscellaneous NP to 0 led to marked increases in coverage of the second, third, and (if applicable) fourth doses, with the absolute change ranging from 9% to 48%, while the first dose coverage was slightly reduced (**Table 2**). When the Miscellaneous NP was fixed, the total doses administered generally increased as the Hospitalization NP increased but did not change substantially as the Breakthrough NP decreased (**Figure 3A1**). The relationship between the Hospitalization NP and Miscellaneous NP was more complex: simultaneously increasing the Hospitalization NP and decreasing the Miscellaneous NP while the Breakthrough NP was fixed led to an increase in doses administered if the Miscellaneous NP was above about 20% of its reference value. When the Miscellaneous NP was decreased further, the impact of the Hospitalization NP became negligible (**Figure 3A2**).

**Figure 3.**
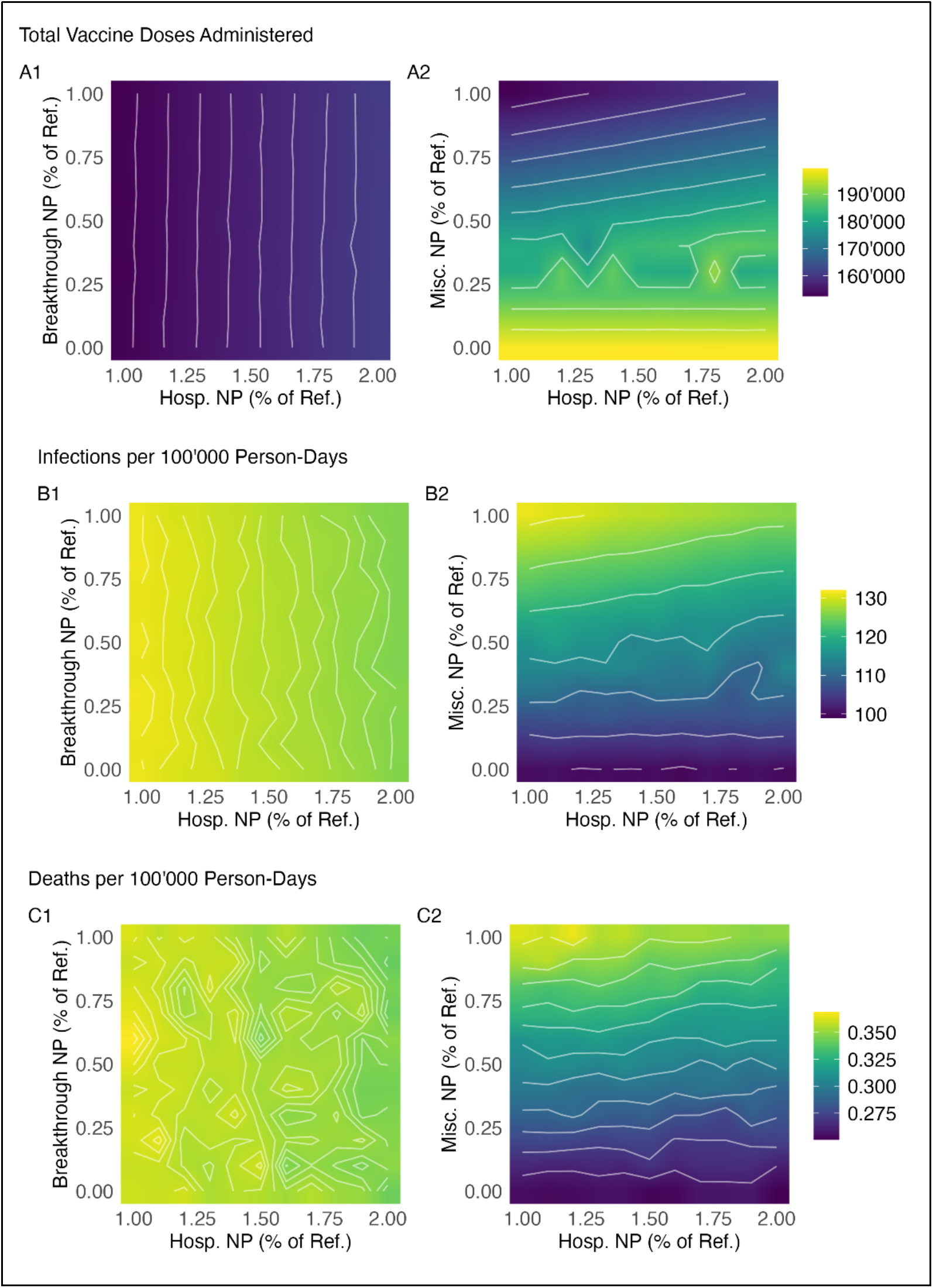
Vaccines administered, incidence rate, and death rate by model scenario. The hospitalization nudge probability (Hosp. NP) was increased from 100% to 200% of its reference value, the breakthrough nudge probability (Breakthrough NP) was decreased from 100% to 0% of its reference value, and the miscellaneous nudge probabilities (Misc. NP) were decreased from 100% to 0% of their reference values, all in increments of 10%. For each parameter combination, the median number of vaccine doses administered per run, the median infection rate per 100’000 person-days, and the median disease-related death rate per 100’000 person-days across 128 runs are displayed.

**Table 2.** Proportion of adult population vaccinated, by dose and age group, for select scenarios. Green indicates higher coverage than the reference level and orange indicates lower (by at least 0.5%). Corresponding 50% simulation intervals are available in the Appendix in Table S3.

| Scenario |  |  | Ages 18 - 49 |  |  | Ages 50 – 64 |  |  |  | Ages 65+ |  |  |  |
| --- | --- | --- | --- | --- | --- | --- | --- | --- | --- | --- | --- | --- | --- |
| *Hospitalization*<br>*Nudge Prob.* | *Breakthrough*<br>*Nudge Prob.* | *Miscellaneous*<br>*Nudge Prob.* | Dose 1 | Dose 2 | Dose 3 | Dose 1 | Dose 2 | Dose 3 | Dose 4 | Dose 1 | Dose 2 | Dose 3 | Dose 4 |
|  |  |  | % | % | % | % | % | % | % | % | % | % | % |
| Ref. | Ref. | Ref. | 74.2 | 61.4 | 18.3 | 74.2 | 61.4 | 36.2 | 7.3 | 89.8 | 78.9 | 51.8 | 17.0 |
| 200% of Ref. | Ref. | Ref. | 77.0 | 64.9 | 22.8 | 77.0 | 64.9 | 40.4 | 8.0 | 91.0 | 81.1 | 55.8 | 18.1 |
| Ref. | 50% of Ref. | Ref. | 74.2 | 61.4 | 18.4 | 74.2 | 61.4 | 36.2 | 7.3 | 89.8 | 78.9 | 51.9 | 17.1 |
| Ref. | 0% of Ref. | Ref. | 74.2 | 61.3 | 18.3 | 74.2 | 61.3 | 36.3 | 7.4 | 89.8 | 78.9 | 52.0 | 17.2 |
| 200% of Ref. | 50% of Ref. | Ref. | 77.0 | 64.9 | 22.9 | 77.0 | 64.9 | 40.5 | 8.0 | 91.0 | 81.1 | 55.8 | 18.1 |
| 200% of Ref. | 0% of Ref. | Ref. | 77.1 | 64.9 | 22.9 | 77.1 | 64.9 | 40.6 | 8.0 | 91.0 | 81.1 | 55.9 | 18.2 |
| Ref. | Ref. | 50% of Ref. | 74.1 | 67.2 | 41.5 | 74.1 | 67.2 | 53.5 | 15.9 | 89.7 | 83.7 | 67.9 | 30.5 |
| Ref. | Ref. | 0% of Ref. | 71.4 | 70.4 | 66.2 | 71.4 | 70.4 | 71.7 | 29.1 | 88.8 | 87.6 | 85.1 | 49.1 |
| 200% of Ref. | Ref. | 50% of Ref. | 77.0 | 70.4 | 45.4 | 77.0 | 70.4 | 57.2 | 16.5 | 90.9 | 85.4 | 70.5 | 31.3 |
| 200% of Ref. | Ref. | 0% of Ref. | 71.4 | 70.4 | 66.2 | 71.4 | 70.4 | 71.6 | 29.1 | 88.8 | 87.6 | 85.1 | 49.1 |

The overall pattern for incidence was the inverse of that for doses: a lower Miscellaneous NP and a higher Hospitalization NP generally led to fewer infections, although the impact of the Hospitalization NP was negligible for the lowest values of the Miscellaneous NP (**Table 3** and **Figure 3B)**. The minimum incidence rate across scenarios was 99.5 infections per 100’000 person-days (50% SI: 97.4, 101.5), corresponding to 31.5 (50% SI: 29.5, 33.7) infections averted per 100’000 person-days compared to the reference scenario; this occurred when the Hospitalization NP was 160% of its reference value and the Miscellaneous NP was 0. The minimum symptomatic case rate across scenarios was 60.3 cases per 100’000 person-days (50% SI: 59.1, 61.1), corresponding to 21.7 (50% SI: 20.8, 22.8) cases averted per 100’000 person-days compared to the reference scenario; this occurred when the Hospitalization NP was doubled and the Miscellaneous NP was 0.

**Table 3.** Overall infection rate and infections averted by end of simulation, for select scenarios. Green indicates fewer infections than the reference level and orange indicates more. Corresponding 50% simulation intervals are available in the Appendix in Table S4.

| Scenario |  |  | Total Infections<br>per 100'000 PD | Infections Averted<br>per 100'000 PD | Percent of<br>Infections Averted | Infections Averted<br>per Addtl. 1'000<br>Doses |
| --- | --- | --- | --- | --- | --- | --- |
| *Hospitalization*<br>*Nudge Prob.* | *Breakthrough*<br>*Nudge Prob.* | *Miscellaneous*<br>*Nudge Prob.* | *n* | *n* | *%* | *n* |
| Reference | Reference | Reference | 131.1 | - | - | - |
| 200% of Reference | Reference | Reference | 125.8 | 5.3 | 4.0 | 391.0 |
| Reference | 50% of Reference | Reference | 131.2 | -0.2 | -0.1 | -232.0 |
| Reference | 0% of Reference | Reference | 131.1 | 0.0 | 0.0 | 1'296.0 |
| 200% of Reference | 50% of Reference | Reference | 125.7 | 5.3 | 4.1 | 388.6 |
| 200% of Reference | 0% of Reference | Reference | 125.8 | 5.2 | 4.0 | 377.9 |
| Reference | Reference | 50% of Reference | 116.0 | 15.0 | 11.4 | 394.1 |
| Reference | Reference | 0% of Reference | 100.1 | 30.9 | 23.6 | 391.8 |
| 200% of Reference | Reference | 50% of Reference | 112.1 | 18.9 | 14.4 | 392.8 |
| 200% of Reference | Reference | 0% of Reference | 99.8 | 31.3 | 23.9 | 396.2 |

**Table 4** and **Figure 3C** demonstrate a similar pattern for deaths, with more noise from model stochasticity. The minimum death rate across scenarios was 0.25 deaths per 100’000 person-days (50% SI: 0.24, 0.27), corresponding to 0.11 deaths averted per 100’000 person-days (50% SI: 0.09, 0.13) compared to the reference scenario; this occurred when the Hospitalization NP was doubled and the Miscellaneous NP was zero.

**Table 4.** Overall death rate and deaths averted by end of simulation, for select scenarios. Green indicates fewer deaths than the reference level and orange indicates more. Corresponding 50% simulation intervals are available in the Appendix in Table S5.

| Scenario |  |  | Total Deaths per<br>100'000 PD | Deaths Averted<br>per 100'000 PD | Percent of Deaths<br>Averted | Deaths Averted<br>per Addtl. 1'000<br>Doses |
| --- | --- | --- | --- | --- | --- | --- |
| *Hospitalization*<br>*Nudge Prob.* | *Breakthrough*<br>*Nudge Prob.* | *Miscellaneous*<br>*Nudge Prob.* | *n* | *n* | *%* | *n* |
| Reference | Reference | Reference | 0.363 | - | - | - |
| 200% of Reference | Reference | Reference | 0.344 | 0.018 | 5.1 | 1.4 |
| Reference | 50% of Reference | Reference | 0.364 | -0.002 | -0.4 | 9.2 |
| Reference | 0% of Reference | Reference | 0.360 | 0.003 | 0.7 | 10.3 |
| 200% of Reference | 50% of Reference | Reference | 0.346 | 0.017 | 4.6 | 1.3 |
| 200% of Reference | 0% of Reference | Reference | 0.342 | 0.020 | 5.6 | 1.4 |
| Reference | Reference | 50% of Reference | 0.305 | 0.058 | 15.9 | 1.5 |
| Reference | Reference | 0% of Reference | 0.254 | 0.108 | 29.8 | 1.4 |
| 200% of Reference | Reference | 50% of Reference | 0.303 | 0.060 | 16.5 | 1.2 |
| 200% of Reference | Reference | 0% of Reference | 0.250 | 0.112 | 31.0 | 1.4 |

**Figure 4** shows the impact of nine selected intervention scenarios on vaccine coverage over time relative to the epidemic curve. Doubling the Hospitalization NP alone (Panel A) affected all vaccine-resistant persons and increased their vaccine uptake relative to the reference scenario across all doses, starting when the hospitalization threshold was typically crossed in late August 2021. As a result, the subsequent January 2022 infection peak was somewhat reduced. Halving or eliminating the Breakthrough NP (Panels B and C) affected only the much smaller population subset who experienced breakthrough infections and caused a nominal increase in their third- and fourth-dose uptake from February 2022 onward. Halving or eliminating the Miscellaneous NP alone (Panels F and G) affected all individuals who received at least one dose and increased their second-, third-, and fourth-dose uptake from February 2021 onward on a much larger scale; as a result, the subsequent August/September 2021 and January 2022 peaks were both more considerably reduced. When the Miscellaneous NP was only halved, the hospitalization threshold was generally still crossed in autumn 2021, so also doubling the Hospitalization NP had an additive effect (Panel H). When the Miscellaneous NP was reduced to zero, however, infections – and, by extension, hospitalizations – were reduced enough that the threshold was no longer crossed at all, making the Hospitalization NP irrelevant (Panel I).

**Figure 4.**
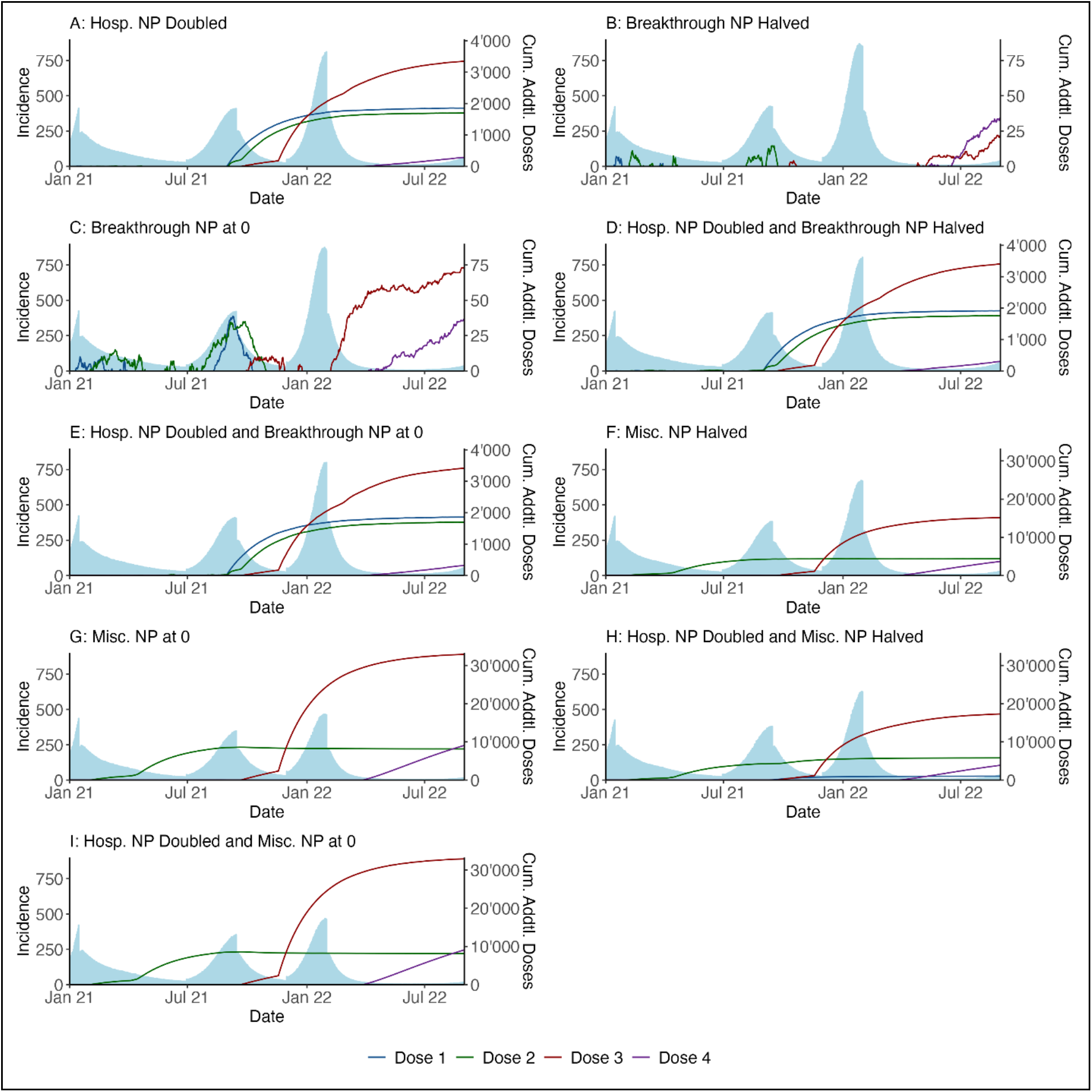
Epidemic curve vs. cumulative vaccines administered over time for select scenarios. The blue regions represent the (median) incident infections by day across the full population of 100’000 nodes. The coloured lines represent the difference between the (median) cumulative number of vaccine doses administered across all age groups in the scenario of interest and the corresponding (median) number from the reference scenario, as of each point in time. Note that the secondary y-axes differ across panels.

### Additional Results

The relative impact of the different intervention scenarios was the same across age targeting approaches, but the interventions acted on a smaller scale when they were limited to those aged 65+ years (**Supplemental Figures S1** and **S2**). Sensitivity analyses of the infection- and vaccine-induced immunity parameters showed that, while the scale of the outbreak varied, the relative impact of the nine selected interventions was similar, except that interventions on the Hospitalization NP became less impactful when the half-life of vaccine induced immunity was increased from 80 days to 140 days or more (**Supplemental Tables S1** and **S2**).

## DISCUSSION

In our study, we used the novel combination of a network-based model of SARS-CoV-2 dynamics and a social-psychological model of COVID-19 vaccination decision-making to investigate which of the pathways between vaccine willingness and resistance might have the greatest influence on vaccine coverage and disease incidence. We found the miscellaneous willing-to-resistant pathway to be the most influential: eliminating the drop-off in vaccine uptake between doses due to community-level miscellaneous factors increased the number of vaccine doses administered by 30.7% and, as a result, decreased the incidence rate by 23.6%. Doubling the probability that a spike in hospitalized prevalence would prompt vaccine hesitant persons to vaccinate increased vaccine uptake by 5.4% and decreased incidence by 4.0%, a smaller but still substantial impact. In contrast, eliminating the probability that a breakthrough infection would prompt a previously vaccine willing person to forgo further vaccination had negligible population-level benefits. Overall, our study suggests that epidemic outcomes are improved when baseline vaccine hesitancy is addressed to reduce the vaccine-naïve population while as many vaccinated persons as possible are kept on the path to subsequent doses, and that optimizing the timing of any vaccine promotion intervention relative to the timing of infection waves – so that the interventions anticipate the case curve instead of reacting to it – is critical.

In our study, interventions affecting persons’ responses to community-level factors were impactful, while those affecting responses to individual-level factors were not, primarily because far more persons were exposed to the former set of interventions than the latter. The miscellaneous willing-to-resistant pathway could affect anyone who received at least one vaccine dose within the model timeframe; it was mostly meant to encompass collective factors, such as widespread false messaging on social media about vaccine-induced female infertility.^21,22^ The hospitalization-related resistant-to-willing-pathway, similarly, could affect anyone who was vaccine resistant when hospitalizations spiked – not just those who were hospitalized themselves. Conversely, the breakthrough-related willing-to-resistant pathway could only affect the small number of fully vaccinated and still vaccine willing individuals who became infected and experienced symptoms despite the layers of protection against both transmission and symptomatic disease provided by the vaccine. Thus, the fundamental difference between changes to the hospitalization or miscellaneous nudge probabilities, which were beneficial on a population level, and changes to the breakthrough nudge probability, which were not, is that the former set of changes had a much wider reach. This finding regarding the importance of scale is consistent with empirical evaluations of vaccine promotion interventions: Athey et al found that the impact of social media advertisements promoting COVID-19 vaccination was small on a per-person basis but in aggregate convinced upwards of 11 million persons.^23^ The distinction we made between individual- and community-level precipitating events is unique to our model. In the Papst et al model of influenza vaccination, individuals based their decisions solely on personal past experiences of infections or vaccine side effects; the two model parameters controlling these events (the vaccine efficacy and the probability of vaccine morbidity) were both relevant to their model predictions.^19^ By considering more varied precipitating events, our study contributes an additional finding: vaccine promotion interventions that focus on experiences shared by wide segments of the population may have more disease prevention potential than those focused on individual experiences with the COVID-19 vaccine in isolation.

Our results also suggest that, due to their better optimized timing, proactive interventions (e.g., changes to the miscellaneous nudge probability) have much more disease prevention potential than interventions that are reactive to naturally occurring transmission events (e.g., changes to the hospitalization and breakthrough nudge probabilities). Lowering the miscellaneous nudge probability meant that as soon as an individual received their first dose, their willingness to receive the next was cemented. As such, not only did changes to the miscellaneous nudge probability cause the largest increase in vaccine coverage, but they also acted the fastest, causing increased dose two uptake as early as February 2021 – the month after the start of the vaccine rollout. These interventions thus affected both infection spikes that occurred within our model timeframe. In contrast, changes to the hospitalization nudge probability could affect the later spike but not the earlier one, which was already well underway by the time the hospitalized prevalence in our model usually crossed its threshold in late August 2021; if the miscellaneous nudge probability was low enough, it could pre-empt interventions on the hospitalization nudge probability altogether by keeping the hospitalized prevalence below its threshold for the full model timeframe. Similarly, the minor increases in third- and fourth-dose uptake caused by changes to the breakthrough nudge probability began very late in the model timeframe, by which point the number of infections that could potentially have been averted was small. These results are consistent with those of other COVID-19 modelling studies that have found the success of a particular vaccination campaign to be highly sensitive to its timing: Gavish et al projected that advancing Israel’s summer 2021 booster campaign by 2 weeks could have halved the number of cases in the subsequent three months.^24^ The models developed by Papst et al and a preceding influenza study by Wells et al both assumed that each annual influenza vaccine rollout concluded before that year’s outbreak began;^19,25^ they therefore did not consider how the rollout’s impact could vary based on timing. Our model’s ability to account for concurrent vaccination and transmission dynamics facilitated our finding that proactively timed interventions – ones that, for example, anticipate increases in hospitalized prevalence instead of reacting to them after the fact – could have a greater impact on epidemic outcomes.

Finally, we found that limiting vaccine promotion interventions to those aged 65 years or older, who are at increased risk of severe outcomes, led to fewer prevention benefits than when either 18- to 64-year-olds or all adults were targeted. This was likely due in part to the relatively small size of the older age group and to their higher reference levels of vaccine willingness, which left less room for intervention-prompted increases. Our study supports findings from previous modelling studies suggesting that vaccine hesitancy should be addressed in younger populations, who are less susceptible to severe disease or death but who play a larger role in transmission and have lower baseline vaccine willingness.^24,26,27^

Our approach has several limitations. First, we only explicitly modelled and tracked three factors that could affect vaccination decisions: vaccine side effects, breakthrough infections, and hospitalized prevalence. To account for all other potential factors, we used a “miscellaneous” willing-to-resistant pathway in our vaccination decision-making model, with receipt of a given vaccine dose used as a proxy for the subsequent, unidentified events that might trigger vaccine resistant attitudes. Without information on the timing of those events, we had to assume that individuals who became vaccine resistant for these other reasons between receiving dose *n* and when they would otherwise have received dose *n + 1* did so immediately after receipt of dose *n*, which may have exaggerated the impact of the miscellaneous nudge due to the timing factors discussed above.

We also did not consider clustering by vaccination type, correlations between a parent’s type and their children’s vaccination status, or the ways that vaccine resistant views might propagate through a network (e.g., how persons might be influenced by breakthrough infections within their household or social network even if they themselves were not a breakthrough case). By assuming that unvaccinated persons were distributed randomly through the network, we may have overestimated the indirect protection they received via vaccinated contacts. This may, in turn, have muted the impact of our interventions, since we did not account for the possibility of the interventions breaking up pockets of vaccine resistance within certain households or neighbourhoods – but as the issue of clustering is complex, it should be explored further by future work on this model.

Another limitation of our model is the uncertainty to which some of our parameters are subject. Due to data limitations, our reference willing-to-resistant nudge probabilities were extrapolated from surveys of vaccine willingness beyond their intended use. Since COVID-19 emerged relatively recently, data on the waning of vaccine-induced and natural immunity is currently limited, and we did not account for heterogeneity in immunity by age, disease severity, or other factors.^28–30^ To address this limitation, we performed sensitivity analyses on the two immunity duration parameters and found that our conclusions about which interventions were most effective generally held.

In conclusion, our findings indicate that addressing community-level factors influencing decision-making may have more disease prevention potential than intervening based on individuals’ own vaccination and infection history, and that attention should be paid to formulating vaccination strategies that accurately predict and pre-empt increases in the case curve. These conclusions were facilitated by a mathematical model that included realistic details of human behaviour based on established results in social psychology, illustrating that models with greater psychological realism can be useful for informing future public health interventions that address barriers to vaccination.

## METHODS

This study used a network-based mathematical model of SARS-CoV-2 transmission, disease progression, and vaccination behaviours in the population of Georgia, USA over a twenty-month period from January 2021 to August 2022 – i.e., the month in which eligibility for the first vaccine dose began to expand in Georgia to the month before the start of the bivalent booster rollout. Our model was built using EpiModelCOVID, a previously validated extension of the EpiModel software platform, which uses the statistical framework of exponential random graph models (ERGMs) to simulate dynamic contact networks.^31^ For this study, we built a social-psychological decision-making model into the vaccination processes within EpiModelCOVID. The model code and software are available on GitHub (https://github.com/EpiModel/COVID-Vax-Decisions).

### Core Model Structure

Our model tracked 100’000 persons (agents) representing a sampled population of the state of Georgia, USA. Agents were assigned an initial age according to Georgia’s age pyramid as of 2020.^32^ They could exit the model population at any time through death (general or disease-specific), while new agents entered the model population exclusively through birth. All modelled agents were members of two distinct, overlapping contact network layers and transmission environments: community and household.

For the community network layer, all contacts (edges) were non-persistent (no duration). Based on the COVIDVu study, we estimated the mean daily degree for this network layer to be 13.8 across all agents and 5.7 across agents aged 65 years or older.^33^ The POLYMOD social mixing study extrapolated to U.S. settings was used to parameterize age mixing, with the within-group contact proportion set at 69% for those aged under 18 years, 81% for 18- to 64-year-olds, and 21% for 65+-year-olds.^34^ The community environment was estimated with an ERGM from which we then simulated at each timestep.

The household network layer was comprised of persistent contacts, lasting from entry into the population to simulation end or death. Each agent was assigned to a household according to an algorithm based on U.S. Census data: 1) 29.2% of households had at least one member aged under 18 years;^35^ 2) 79.1% of households had at least one 18- to 64-year-old;^35^ 3) 31.4% of households had at least one 65+-year-old;^35^ 4) the average household had 2.7 persons;^36^ 5) every household with a child also had at least one adult; and 6) 97.9% of children had an 18- to 64-year-old in their household.^37^ We took this approach due to the lack of recent social mixing data for children in U.S. settings. Household edges were specified such that each household was fully saturated and each edge was within a single household. Community and household contacts were subsequently combined to create a multi-layer dynamic network.

Our model represented the natural history of COVID-19 using a SEIRS framework. Susceptible agents could stochastically transition to the exposed state upon contact with an infected person (i.e., a discordant contact). The daily probability of infection given a discordant contact depended on the vaccination status of the susceptible agent, the symptom status of the infectious agent, and whether the contact was household- or community-level. Newly infected agents were stochastically assigned to either an asymptomatic or symptomatic clinical pathway, with a subset of symptomatic agents subsequently designated for hospitalization. The probabilities of symptoms and hospitalization both depended on age and vaccination status. Agents in the hospitalized state experienced a higher age-specific mortality rate than those in other states. Once recovered, agents stochastically re-entered the susceptible state, where they could be reinfected. Parameters defining the model’s disease progression, transmission, and clinical epidemiology (**Table 1**) were either drawn from existing literature or calibrated as described below.

### Vaccination Decision-Making Process

Each adult agent was assigned a binary vaccination “type”– resistant or willing – such that the prevalence of vaccine willingness by age group at the start of the vaccine rollout matched the empirical distribution in late 2020.^38,39^ The vaccination decision-making process for agents aged under 18 years was not explicitly modelled, given fundamental differences in this process for minors versus adults. Instead, children received vaccine doses according to age-specific rates.

In accordance with the law of inertia in decision-making,^18^ agents maintained their initial attitude toward vaccination until an adverse event (“nudge”) prompted them to change – meaning disease outcomes could affect vaccination behaviour, which in turn affected future disease outcomes. Four such nudges were considered: 1) experiencing vaccine side effects could prompt a vaccine willing individual (who had received at least one dose) to become vaccine resistant; 2) experiencing an infection while fully vaccinated (a “breakthrough infection”) could prompt a vaccine willing individual (who had received at least two doses) to lose trust in the vaccine and become vaccine resistant; 3) increased hospitalized prevalence in the population could prompt any vaccine resistant individual to grow more concerned about the spread of COVID-19 and become vaccine willing; and 4) an additional willing-to-resistant pathway covered all other reasons for developing vaccine resistant attitudes (among individuals who had received at least one dose), such as social conformity or media influences. For this fourth miscellaneous pathway, the timing of an individual’s last vaccine dose was used as a proxy for the timing of the unknown precipitating event prompting resistance towards future doses.

To parameterize the first two nudges, we identified: 1) the odds ratio comparing the likelihood of booster willingness for those who had versus had not missed work due to side effects from the primary vaccination series;^40^ and 2) the odds ratio comparing the likelihood of booster willingness for those who had received the primary vaccination series and had versus had not been subsequently infected.^41^ We converted these odds ratios to one-time probabilities of being “nudged” towards vaccine resistance. For the third nudge, we estimated the probability that a vaccine-eligible adult in Georgia was convinced to vaccinate by an increase in hospitalized COVID-19 prevalence between July and September 2021, using the finding that 38% of that period’s surveyed late adopters were motivated by concern about local hospitalizations.^9^ The probabilities of behaviour change associated with the fourth nudge were treated as free parameters in the calibration process.

At any timestep, agents could stochastically undergo vaccination if they were not currently symptomatic, had not tested positive in the last two weeks, were vaccine willing (for adult agents), and were currently eligible for their next dose based on their age group and vaccination history. Vaccination reduced the risk of disease acquisition, the risk of progression to symptomatic disease, and the risk of eventual hospitalization. Vaccine immunity waned over time following an exponential decay with a half-life of 80 days.^42^

### Calibration

The per-act infection probability was calibrated so that the simulated number of incident infections by month matched the confirmed case counts reported by the Georgia Department of Public Health (GDPH),^43^ adjusted to account for underreporting^44^ and scaled to a population of 100’000. To account for time-varying coverage of non-pharmaceutical interventions and the introduction of new variants, this per-act infection probability was boosted by 34% for two periods of increased transmission and suppressed by 40% for three periods of decreased transmission. The age-specific hospitalization proportions and disease-related mortality multiplier were calibrated so that the resultant COVID-19 related hospital admissions and deaths by month matched GDPH reports.^43^ Finally, age- and dose-specific vaccination rates and the nudge probabilities for the miscellaneous willing-to-resistant pathway were calibrated so that the resultant vaccine coverage by age, dose, and month matched the levels reported by the CDC for Georgia.^4^

### Intervention Scenarios

We compared our calibrated model to counterfactual scenarios that explored hypothetical interventions on three of the four nudges in our model. These interventions did not affect the precipitating events, but rather the agents’ responses to them. Specifically, we 1) kept the probability that vaccine side effects would prompt a vaccine willing-to-resistant switch constant across all scenarios; 2) decreased the probability that a breakthrough infection would prompt a willing-to-resistant switch (the Breakthrough Nudge Probability or Breakthrough NP) to as little as zero; 3) increased the probability that a spike in hospitalizations would prompt a resistant-to-willing switch (the Hospitalization Nudge Probability or Hospitalization NP) to as much as two times the reference value; and 4) decreased the probability of a vaccine willing-to-resistant switch related to something other than side effects or breakthrough infections (the Miscellaneous Nudge Probability or Miscellaneous NP) to as little as zero. We then also explored the impact of applying these changes to only older adults (65+-year-olds) or only younger adults (18- to 64-year-olds) instead of all adult agents.

Since data on the duration of COVID-19 immunity was limited, we performed sensitivity analyses on two particularly uncertain parameters: the average duration of natural immunity to infection after recovery and the half-life of vaccine-induced immunity.

### Model Output

For each model scenario, we tracked the incidence rate of SARS-CoV-2 infections, the COVID-related death rate, and the per-dose vaccine coverage by age group over time. The median and 50% simulation interval (SI) of each output across 128 simulations per scenario were reported.

## Data Availability

The model code and software are available on GitHub at https://github.com/EpiModel/COVID-Vax-Decisions.

