## Supplemental Appendix for "Modelling the Interplay between Responsive Individual Vaccination Decisions and the Spread of SARS-CoV-2"

### 1 INTRODUCTION

This supplementary technical appendix describes in further detail the structure, parameterization, and analysis of the mathematical model used in the accompanying paper.

#### 1.1 Model Framework

The mathematical models for SARS-CoV-2 transmission dynamics presented in this study were network-based transmission models in which uniquely identifiable contact dyads were simulated and tracked over time. This contact structure was represented using exponential-family random graph models (ERGMs), described in Section 2. On top of this dynamic network simulation, the epidemic model represented demography (entries, exits, and aging), interhost epidemiology (disease transmission), intra-host epidemiology (disease progression), and clinical epidemiology (disease diagnosis and prevention interventions). Individual attributes related to these processes were stored and updated in discrete time over the course of each epidemic simulation.

#### 1.2 Model Software

The models in this study were programmed in the R and C++ software languages using the *EpiModel* [http://epimodel.org/] software platform for epidemic modelling, which was developed for simulating complex network-based mathematical models of infectious diseases.[1] *EpiModel* depends on *Statnet* [http://statnet.org/], a suite of software in R for the representation, visualization, and statistical analysis of complex network data.[2]

*EpiModel* allows for a modular expansion of its built-in modelling tools to address novel research questions. Jenness et al. developed a set of extension modules into a software package called *EpiModelCOVID* for use in a prior SARS-CoV-2 modelling study,[3] and we adapted these modules to address our research questions. The updated software and the scripts used in the execution of these models are available for download in two GitHub repositories:

1. [https://github.com/EpiModel/EpiModelCOVID] contains the general extension software package. Installing this using the instructions on the repository homepage will also load in EpiModel and the other dependencies. We use a branching repository architecture on GitHub; the branch of the repository associated with this research project is Vax-Decisions.
2. [https://github.com/EpiModel/COVID-Vax-Decisions] contains the scripts to execute the models and to run the analyses provided in the manuscript.

##### 1.3 Core Model Specification

We started with a network of 100’000 agents aged 0 to 100 to represent the larger population of the state of Georgia. The population size was allowed to increase and decrease with arrivals (new births) and departures related to general or disease-specific mortality. Further details on the demography are provided in Section 3. We used a two-stage simulation framework, first calibrating the model to total SARS-CoV-2 incidence, COVID-related hospital admissions, COVID-related deaths, and age- and dose-specific vaccine coverage (Stage 1), then simulating the reference and counterfactual intervention scenarios for a period of 608 days corresponding to 01 January 2021 to 31 August 2022 (Stage 2). The time unit used throughout the simulations was one day. Unless otherwise noted, all rate-based parameters in this appendix are to be interpreted as the rate per day and all duration-based parameters are to be interpreted as the duration in days.

#### 2 CONTACT NETWORKS

We modelled two distinct contact networks: household and community. Here we first describe the methods conceptually, including the parameters used to guide the model and their derivation, and then present the formal statistical modelling methods.

##### 2.1 Conceptual Representation of Community Contact Network

Our modelling methods aimed to preserve certain features of the cross-sectional and dynamic network structure observed in our data sources, within the context of changing population size (due to births and deaths from the population) and changing composition by attributes such as age.

The network features that we aimed to preserve were as follows:

1. The mean degree (mean number of persons contacted in one day) across the full population.
2. The mean degree (mean number of persons contacted in one day) across the population of individuals aged 65 years and over.
3. The level of assortative mixing within each of the following age groups: under 18 years, at least 18 and under 65 years, and 65 years and over.

##### 2.1.1 Overall Mean Degree for Community Network

The overall cross-sectional mean degree across the population for the community network layer was estimated at 13.8, based on the results of the COVIDVu study, which surveyed a probability sample of 3'112 U.S. households in spring 2021 and found 13.8 to be participants' mean number of daily contacts across all non-household settings (work, school, and other).[4]

##### 2.1.2 Heterogeneity in Mean Degrees for Community Network

Also based on the results of the COVIDVu study, the mean degree for nodes aged 65 years or older was estimated to be about 41% of the overall mean degree, or about 5.7.[4] (Mean degree targets for the younger two age groups, under 18 years and 18 to 64 years, were not specified in the network estimation process, but after this process was complete, the mean degrees for these two groups worked out to about 17.1 and 14.5 contacts per day, respectively, across 1'000 test networks.)

##### 2.1.3 Mixing by Age in Community Network

Based on the results of the POLYMOD social mixing study extrapolated to U.S. settings, we estimated that 69% of edges for those aged under 18 years, 81% of edges for those aged at least 18 and under 65 years, and 21% of edges for the those aged 65 years and over should be within-group.[5]

#### 2.2 Statistical Representation of Community Contact Network

Exponential-family random graph models (ERGMs) and their dynamic extension temporal ERGMs (TERGMs) provide a foundation for statistically principled simulation of local and global network structure given a set of target statistics from empirical data. Community contacts were assumed to last for one day – meaning each node could receive a new set of contacts at each daily time step, without regard for which nodes they were previously connected to – and were therefore modelled using cross-sectional ERGMs.[6] Formally, our statistical model for community contact dynamics can be represented using the following equation for the conditional log odds (logits) of relational existence at time *t*:

$$logit\left(P\left(Y_{ij,t} = 1 \middle| Y_{ij,t}^{C}\right)\right) = \theta'\partial\left(g(y)\right)$$

where:

- $Y_{ij,t}$ = the relational status of persons *i* and *j* at time *t* (1 = in contact, 0 = not).
- $Y^{C}_{ij,t}$ = the network complement of *i,j* at time *t*, i.e., all relations in the network other than *i,j*.
- $g(y)$ = vector of network statistics for the model (the empirical statistics defined in the previous section).
- $\partial\big(g(y)\big)$ = the change in $g(y)$ when $Y_{ij}$ is toggled from 0 to 1.
- $\theta$ = vector of parameters in the model.

The recursive dependence among the relationships rendered the model impossible to evaluate using standard techniques; we used MCMC to obtain the maximum likelihood estimates for the $\theta$ vector given the $g(y)$ vector.

Our method of converting the statistics laid out in Section 2.1 into our fully specified network model consisted of the following steps:

1. Construct a cross-sectional network of 100’000 persons with no relationships.
2. Assign ages based on Georgia’s age pyramid.
3. Calculate the target statistics (i.e., the expected count of each statistic at any given moment in time) associated with the terms in the existence model for community contacts.
4. Estimate the coefficients for the existence model that represent the maximum likelihood estimates for the expected cross-sectional network structure.

These steps occurred within the *EpiModel* software and used the ERGM methods therein. They were completed efficiently by use of an approximation in Step 4.[7] During the subsequent model simulation, we used the method of Krivitsky to adjust the coefficient for the edges term in each model at each time step, to preserve the same expected mean degree (contacts per person) over time in the face of changing network size and nodal composition.[8] At all stages of the project, simulated partnership networks were checked to ensure that they indeed retained the expected cross-sectional structure throughout the simulations.

##### 2.3 Algorithmic Approach to Household Network

Due to the lack of recent (pandemic-era) social mixing data for children in U.S. settings, it was not feasible to specify the target statistics required by the ERGM methods described above for the household network. Instead, each agent was algorithmically assigned to a household according to the following six rules:

1. 29.2% of households had at least one member aged under 18 years.
2. 79.1% of household had at least one member aged at least 18 and under 65 years.
3. 31.4% of households had at least one member aged 65 years or older.
4. The average household had about 2.7 persons.
5. Every household with a child also had at least one adult.
6. 97.9% of individuals aged under 18 years had an individual aged at least 18 and under 65 years in their household.

Rules 1 through 3 were based on Census data about the composition of U.S. households;[9] Rule 4 was based on Census data for Georgia specifically;[10] Rule 5 was an assumption; and Rule 6 was a simplified interpretation of the Census finding that 2.1% of U.S. children lived with at least one grandparent and without a parent as of 2021.[11]

Agents were then mapped to households using an algorithm with the following steps:

1. The total number of households was calculated by dividing the number of agents by the average household size.
2. 29.2% of households were randomly selected and one child was assigned to each of the selected households. All remaining children were then randomly allocated across the selected households.
3. A subset of the households with children was selected and designated as needing a younger adult (aged between 18 and 64 years), such that the designated households together contained 97.9% of all children.
4. Additional households without children were randomly selected and designated as needing a younger adult, such that 79.1% of households were designated in total (including those from Step 3). One younger adult was assigned to each of these households and all remaining younger adults were randomly allocated across them.
5. All households without a younger adult were designated as needing an elderly person (aged 65+ years). (This included empty households as well as households that so far only contained children). Additional households were randomly selected (from among the households with a younger adult) and designated as needing an elderly person such that 31.4% of households were designated in total. One elderly person was assigned to each of these households and all remaining elderly persons were randomly allocated across them.

Once every individual had been allocated to a household, a list of household pairings was constructed such that each household was fully saturated and each edge was within a single household. These pairings were kept constant over time, lasting from simulation start or birth (whichever came first) to simulation end or death (whichever came first). They were subsequently combined with the community pairings to create the multi-layer dynamic network used in the final simulations.

### 3 DEMOGRAPHY

In this model, there were three demographic processes: entries, exits, and aging. Entries were conceptualized as births and exits as deaths; immigration into and emigration out of the population of interest were not explicitly accounted for.

#### 3.1 Arrivals

All persons entered the network at age 0. The birth rate (per person per day) was set at the average of the age-specific general mortality rates to keep the overall network size relatively stable. At each time step, the exact number of individuals entering the population was simulated by drawing from a Poisson distribution with the distribution mean set at the per-person birth rate multiplied by the current population size.

#### 3.2 Initialization of Attributes

Persons born into the population were assigned the following attributes:

- **Age.** As noted above, all incoming nodes were assigned an initial age of 0, which incrementally grew in daily time steps.
- **Household.** To maintain the desired proportion of households with children, each newly born individual was randomly assigned to a household that already had a member aged under 18 years.
- **Disease status.** All new births entered the population in the susceptible state.
- **Vaccine status.** All new births entered the population unvaccinated.
- **Vaccinator type.** Only adult agents in the model were assigned a vaccinator type, so new births had their vaccinator type temporarily designated as "NA."

#### 3.3 Departures from the Network

All exits due to mortality were modelled stochastically. Departures included both natural (non-COVID) and disease-induced mortality causes. Background mortality rates were based on Georgia age-specific all-cause mortality rates from the OASIS Mortality Web Query.[12] These yearly rates were transformed into daily risks. For modelled individuals in the COVID-19 hospitalized disease state, a calibrated mortality multiplier was applied to their base mortality risk. (Thus, we assumed that no one died a COVID-related death without being hospitalized first, and that all deaths from the COVID-19 hospitalized state could be attributed to COVID-19.) Mortality was then applied to all persons within the population at each time step stochastically by drawing from a Bernoulli distribution for each person with a probability parameter corresponding to their age- and disease state-specific risk of death.

#### 3.4 Aging

The aging process in the population was linear by time step for all persons. Evolving age impacted background mortality, age-based mixing in forming new partnerships, and other features of the epidemic model described below. Additionally, upon reaching the age of 18 years, agents were assigned a binary "vaccinator type" attribute – resistant or willing – based on the reported proportion of 18- to 49-year-olds who were vaccine willing at the start of the vaccine rollout.[13]

### 4 INTRAHOST EPIDEMIOLOGY

Intra-host epidemiology in our model included features related to the natural disease progression within COVID-19 infected persons, which we represented using a modified SEIRS framework. At simulation start, all individuals were either susceptible or exposed, and all new births entered the population in the susceptible state. Once exposed via contact with an infectious person, newly infected individuals were stochastically assigned to either the asymptomatic or symptomatic clinical pathway, with the probability of symptoms dependent on age and vaccination status. Symptomatic individuals progressed from the exposed state through the infectious pre-symptomatic state to the infectious symptomatic state; from the infectious symptomatic state, individuals could stochastically enter the hospitalized state before recovery or directly enter the recovered state, with the probability of hospitalization also dependent on age and vaccination status. Asymptomatic individuals progressed directly from the exposed state to the asymptomatic state and then on to the recovered state. Once recovered, individuals stochastically re-entered the susceptible population, where they could be reinfected. Overall, individuals in the infectious pre-symptomatic, infectious symptomatic, or asymptomatic states were considered infectious; individuals in the hospitalized state were assumed to be effectively isolated and therefore unable to transmit their infection.

#### 5 INTERHOST EPIDEMIOLOGY

Interhost epidemiological processes represented the SARS-CoV-2 transmission process within the model. The model's SEIRS framework allowed susceptible agents to stochastically transition from the susceptible to the exposed state (i.e., to become infected) upon contact with an infected person (i.e., a discordant contact). The per-act probability of infection given a discordant contact depended on the vaccination status of the susceptible agent, the symptom status of the infectious agent, and whether the contact was household- or community-level.

Specifically, asymptomatic infected persons had 50% the transmission potential of those with symptomatic infections.[14] The daily act rate for the community network layer was assumed to be one (so that the per-act infection probability was equivalent to the daily infection probability for this layer); the daily act rate for the household network layer was calculated such that the total probability of infection from a household contact across a five-day infectious period approximately matched the secondary attack rate reported by a household transmission study.[15] As a result, household contacts had about three times the per-day transmission potential of community contacts, accounting for increased opportunity for transmission via what were assumed to be closer, more sustained contacts.

#### 6 CLINICAL EPIDEMIOLOGY

The two main clinical epidemiological processes included in the model were testing & diagnosis and vaccination.

##### 6.1 Testing & Diagnosis

Both uninfected and infected persons in our model were exposed to regular COVID-19 testing. Testing rates were higher for symptomatic infected persons than for persons in all other disease states (10% per day vs. 1% per day).[3] The main impact of a positive diagnosis in the model was that persons were not considered eligible for vaccination for the following 10 days. Interventions such as case isolation were not explicitly accounted for in the model, although their real-world effects were reflected in the calibrated, time-varying transmission probabilities.

#### 6.2 Vaccination

At simulation start or upon reaching the age of 18 years, whichever came first, each adult agent was assigned a binary “vaccinator type” attribute – resistant or willing – such that the prevalence of vaccine willingness by age group at the start of the vaccine rollout matched the empirical distribution measured in late 2020.[13,16] (Agents aged under 18 years simply received vaccine doses according to age group-specific but otherwise homogenous rates.) Agents maintained their initial attitude toward vaccination until one of the following events prompted them to reconsider:

1. **Vaccine side effects:** At each time step, we applied vaccine side effects stochastically by drawing from a Bernoulli distribution for newly vaccinated person with a probability parameter corresponding to their dose-specific risk of side effects.[17] (“Side effects” were defined as any reaction to a vaccine severe enough to cause an individual to miss work.) At the subsequent timestep, those individuals who had been newly selected for side effects could stochastically transition from vaccine willing to vaccine resistant with a certain probability. This “nudge probability” was estimated from the odds ratio comparing the odds of booster willingness for those who had versus had not missed work due to side effects from the primary vaccination series by converting it to a risk difference.[17]
2. **Breakthrough infections:** If an individual who was fully vaccinated and (if applicable) up to date on their booster vaccinations became infected and began experiencing symptoms, then at the time step in which their symptoms started, they could stochastically transition from vaccine willing to vaccine resistant with a certain probability. This “nudge probability” was estimated from the odds ratio comparing the odds of booster willingness for those who had received the primary vaccination series and had versus had not been subsequently infected by converting it to a risk difference.[18]
3. **Hospitalization spike:** Hospitalized prevalence in the model crossing 36 cases per 100’000 persons for the first time since the start of the vaccine rollout could prompt vaccine resistant individuals to become willing due to heightened concern about the effects of the spread of COVID-19. In order to calculate the “nudge probability” for this pathway, we estimated the proportion of unvaccinated, vaccine-eligible adults in Georgia who were convinced to vaccinate by an increase in hospitalized COVID-19 prevalence between July and September 2021 (during the Delta wave), using one survey’s finding that 38% of that period’s late adopters were motivated by concern about local hospital capacity.[19] Since the percentage of hospital beds in Georgia occupied by COVID-19 patients increased from about 2% to about 29% during this period, we treated 20% as the threshold of interest;

20% of Georgia's 19'747 hospital beds translated to 3'949 hospitalized COVID patients out of Georgia's approximately 10.8 million people, or about 36 hospitalized COVID cases per 100'000 persons.[20]

4. **Other:** We also included an additional willing-to-resistant pathway to cover all other reasons for developing vaccine resistant attitudes – such as social conformity, friends' and family members' experiences with vaccines and COVID-19, or the spread of information via news outlets and social media. For this pathway, receipt of a vaccine dose was used as the precipitating event that could, with a certain probability, trigger vaccine resistant views towards future doses (making vaccination a proxy for the actual, unidentified trigger). The nudge probabilities for this pathway were treated as free parameters in the calibration process for vaccine coverage.

The vaccination rollout in the model proceeded by age group (0 to 4, 5 to 17, 18 to 49, 50 to 64, and 65+ years), with the oldest group becoming eligible for the first dose on Day 11 of each simulation (corresponding to 11 January 2021, when everyone aged 65 years or over became eligible for the first dose in Georgia).[21] Individuals became eligible for their second dose 21 days after receiving their first;[22] they became eligible for their first booster once (1) it had been rolled out to their age group and (2) six months had passed since they received their second dose;[23] they then became eligible for their second booster once (1) it had been rolled out to their age group and (2) four months had passed since they received their first booster.[24] At any given timestep, individuals could stochastically undergo vaccination if (1) they were not currently symptomatic and had not tested positive in the last two weeks, (2) they were vaccine willing (for adult agents), and (3) they were currently eligible for their next dose based on their age group and vaccination history.

Vaccination reduced the risk of disease acquisition, the risk of progression to symptomatic disease among those infected, and the risk of eventual hospitalization among those with symptoms. These effects were dose specific. Vaccine immunity waned over time following an exponential decay pattern with a half-life of 80 days.[25]

#### 7 MODEL CALIBRATION

Each model simulation was initialized with 1'600 persons in the exposed (infected but not infectious) state. Simulations were each run for an initial period of 180 daily time steps (corresponding to 5 July 2020 through 31 December 2020) for calibration purposes; the results of this calibration period were then used as the initial conditions for the simulation of the timeframe of interest: a period of 608 days, corresponding to 1 January 2021 to 31 August 2022.

The free parameters in the calibration process were as follows:

1. The base per-act infection probability.
2. The per-act infection probability during times of increased transmission.
3. The per-act infection probability during times of decreased transmission.
4. The start and end time steps for each period of increased transmission (2) or decreased transmission (3).
5. A scaling factor for the base age-specific probability of hospitalization.
6. The multiplier applied to the probability of hospitalization during the period of increased disease severity (the Delta wave).
7. The multiplier applied to the probability of hospitalization during the period of decreased disease severity (the Omicron wave).
8. The start and end time steps for the period of increased severity and the period of decreased severity.
9. The daily vaccination rates by age group and dose.
10. The “miscellaneous nudge probability” by age group and dose (i.e., the probability that someone in each age group would cease to vaccinate after receiving a given dose due to a reason other than side effects or breakthrough infections.)
11. The disease-specific mortality multiplier applied to the general mortality rate for persons in the model hospitalized due to COVID-19.

The calibration targets were as follows:

1. The monthly confirmed case counts reported by the Georgia Department of Public Health, multiplied by five to account for underreporting and scaled to a population of 100’000.[26,27]
2. The monthly confirmed COVID-19 related hospitalizations reported by the Georgia Department of Public Health, scaled to a population of 100’000.[26]
3. The monthly confirmed COVID-19 related deaths reported by the Georgia Department of Public Health, scaled to a population of 100’000.[26]
4. The vaccine coverage levels by age, dose, and month reported by the CDC for Georgia.[28]

#### SUPPLEMENTAL FIGURES & TABLES

**Table S1. Incident infections per 100’000 person-days for select scenarios and varying durations of naturally induced immunity.** The average duration of immunity after infection was varied from its reference value (300 days) in increments of 30 days. For each scenario and immunity duration value, the median number of infections per 100’000 person-days across 128 runs is reported below.

| **Scenario** | | | **Average Duration of Naturally Induced Immunity** | | | | | |
|---|---|---|---|---|---|---|---|---|
| *Hosp. Nudge Prob.* | *Breakthrough Nudge Prob.* | *Misc. Nudge Prob.* | *270 Days* | *300 Days (Ref.)* | *330 Days* | *360 Days* | *390 Days* | *420 Days* |
| Reference | Reference | Reference | 146.3 | 130.9 | 115.5 | 103.1 | 90.9 | 84.3 |
| 200% of Reference | Reference | Reference | 141.1 | 125.9 | 105.8 | 98.5 | 86.5 | 77.5 |
| Reference | 0% of Reference | Reference | 146.5 | 131.1 | 115.5 | 102.7 | 90.7 | 82.4 |
| 200% of Reference | 0% of Reference | Reference | 141.4 | 125.6 | 111.3 | 99.3 | 86.5 | 77.3 |
| Reference | Reference | 0% of Reference | 114.9 | 100.2 | 83.3 | 68.3 | 58.2 | 58.2 |
| 200% of Reference | Reference | 0% of Reference | 114.6 | 100.1 | 80.8 | 65.8 | 56.7 | 56.5 |

**Table S2. Incident infections per 100'000 person-days for select scenarios and varying half-lives of vaccine-induced immunity.** The half-life controlling the exponential decay of immunity after vaccination was varied from its reference value (80 days) in increments of 30 days. For each scenario and immunity duration value, the median number of infections per 100'000 person-days across 128 runs is reported below.

| **Scenario** | | | **Half-Life of Vaccine Induced Immunity** | | | | | |
|---|---|---|---|---|---|---|---|---|
| *Hosp. Nudge Prob.* | *Breakthrough Nudge Prob.* | *Misc. Nudge Prob.* | *50 Days* | *80 Days (Ref.)* | *110 Days* | *140 Days* | *170 Days* | *200 Days* |
| Reference | Reference | Reference | 149.1 | 131.2 | 116.2 | 105.0 | 96.8 | 92.3 |
| 200% of Reference | Reference | Reference | 146.0 | 126.2 | 109.9 | 105.2 | 97.0 | 92.2 |
| Reference | 0% of Reference | Reference | 149.1 | 131.2 | 115.6 | 104.5 | 96.6 | 92.2 |
| 200% of Reference | 0% of Reference | Reference | 146.0 | 126.1 | 109.7 | 104.0 | 97.1 | 92.1 |
| Reference | Reference | 0% of Reference | 128.6 | 100.1 | 87.0 | 80.8 | 78.8 | 77.3 |
| 200% of Reference | Reference | 0% of Reference | 126.7 | 99.8 | 86.6 | 81.0 | 78.5 | 76.5 |

**Table S3. Proportion of adult population vaccinated by dose and age group for select scenarios, with 50% simulation intervals.** For each scenario, the median, 25th percentile, and 75th percentile of each outcome across 128 runs are reported below. This table corresponds to Table 2 in the main results section, in which only the medians were reported for the sake of conciseness. (The percentages below represent coverage among the full population in each age group, not only the eligible population. Results for 18- to 49-year-olds and 50- to 64-year-olds were combined for the first two doses but treated separately for booster doses to match CDC reports and because those under 50 years of age were not eligible for the fourth dose.)

| **Scenario** | | | **Ages 18 - 49** | | | **Ages 50 – 64** | | | | **Ages 65+** | | | |
|---|---|---|---|---|---|---|---|---|---|---|---|---|---|
| *Hosp. Nudge Prob.* | *Breakthrough Nudge Prob.* | *Misc. Nudge Prob.* | Dose 1 % | Dose 2 % | Dose 3 % | Dose 1 % | Dose 2 % | Dose 3 % | Dose 4 % | Dose 1 % | Dose 2 % | Dose 3 % | Dose 4 % |
| Ref. | Ref. | Ref. | 74.2 (74.1, 74.3) | 61.4 (61.2, 61.5) | 18.3 (18.1, 18.4) | 74.2 (74.1, 74.3) | 61.4 (61.2, 61.5) | 36.2 (35.9, 36.4) | 7.3 (7.2, 7.4) | 89.8 (89.7, 90.0) | 78.9 (78.7, 79.1) | 51.8 (51.5, 52.2) | 17.0 (16.8, 17.3) |
| 200% of Ref. | Ref. | Ref. | 77.0 (76.9, 77.1) | 64.9 (64.8, 65.0) | 22.8 (22.7, 22.9) | 77.0 (76.9, 77.1) | 64.9 (64.8, 65.0) | 40.4 (40.1, 40.7) | 8.0 (7.9, 8.1) | 91.0 (90.8, 91.2) | 81.1 (80.9, 81.3) | 55.8 (55.4, 56.2) | 18.1 (17.8, 18.3) |
| Ref. | 50% of Ref. | Ref. | 74.2 (74.1, 74.3) | 61.4 (61.2, 61.5) | 18.4 (18.3, 18.5) | 74.2 (74.1, 74.3) | 61.4 (61.2, 61.5) | 36.2 (35.9, 36.6) | 7.3 (7.2, 7.5) | 89.8 (89.7, 90.0) | 78.9 (78.7, 79.1) | 51.9 (51.5, 52.1) | 17.1 (16.8, 17.3) |
| Ref. | 0% of Ref. | Ref. | 74.2 (74.1, 74.3) | 61.3 (61.2, 61.4) | 18.3 (18.2, 18.5) | 74.2 (74.1, 74.3) | 61.3 (61.2, 61.4) | 36.3 (35.9, 36.6) | 7.4 (7.2, 7.5) | 89.8 (89.7, 90.0) | 78.9 (78.7, 79.1) | 52.0 (51.5, 52.3) | 17.2 (16.9, 17.4) |
| 200% of Ref. | 50% of Ref. | Ref. | 77.0 (76.9, 77.2) | 64.9 (64.8, 65.0) | 22.9 (22.7, 23.1) | 77.0 (76.9, 77.2) | 64.9 (64.8, 65.0) | 40.5 (40.2, 40.8) | 8.0 (7.8, 8.1) | 91.0 (90.8, 91.1) | 81.1 (80.8, 81.3) | 55.8 (55.5, 56.1) | 18.1 (17.9, 18.3) |
| 200% of Ref. | 0% of Ref. | Ref. | 77.1 (76.9, 77.2) | 64.9 (64.8, 65.1) | 22.9 (22.8, 23.1) | 77.1 (76.9, 77.2) | 64.9 (64.8, 65.1) | 40.6 (40.3, 40.9) | 8.0 (7.8, 8.2) | 91.0 (90.8, 91.1) | 81.1 (80.9, 81.3) | 55.9 (55.5, 56.2) | 18.2 (17.9, 18.5) |
| Ref. | Ref. | 50% of Ref. | 74.1 (71.6, 74.3) | 67.2 (64.3, 67.4) | 41.5 (38.1, 41.7) | 74.1 (71.6, 74.3) | 67.2 (64.3, 67.4) | 53.5 (50.7, 54.0) | 15.9 (15.6, 16.2) | 89.7 (89.0, 89.9) | 83.7 (82.5, 83.9) | 67.9 (65.9, 68.3) | 30.5 (30.1, 30.8) |
| Ref. | Ref. | 0% of Ref. | 71.4 (71.3, 72.0) | 70.4 (70.2, 70.9) | 66.2 (66.0, 66.6) | 71.4 (71.3, 72.0) | 70.4 (70.2, 70.9) | 71.7 (71.4, 72.4) | 29.1 (28.8, 29.3) | 88.8 (88.6, 89.2) | 87.6 (87.5, 88.2) | 85.1 (84.9, 85.5) | 49.1 (48.8, 49.3) |
| 200% of Ref. | Ref. | 50% of Ref. | 77.0 (75.4, 77.1) | 70.4 (68.7, 70.6) | 45.4 (41.6, 45.7) | 77.0 (75.4, 77.1) | 70.4 (68.7, 70.6) | 57.2 (53.2, 57.5) | 16.5 (16.1, 16.8) | 90.9 (90.2, 91.0) | 85.4 (84.4, 85.6) | 70.5 (67.8, 70.9) | 31.3 (30.5, 31.7) |
| 200% of Ref. | Ref. | 0% of Ref. | 71.4 (71.3, 71.7) | 70.4 (70.2, 70.6) | 66.2 (66.0, 66.4) | 71.4 (71.3, 71.7) | 70.4 (70.2, 70.6) | 71.6 (71.3, 71.9) | 29.1 (28.9, 29.3) | 88.8 (88.5, 89.1) | 87.6 (87.4, 88.0) | 85.1 (84.8, 85.4) | 49.1 (48.8, 49.4) |

**Table S4. Overall infection rate and infections averted for select scenarios, with 50% simulation intervals.** For each scenario, the median, 25th percentile, and 75th percentile of each outcome across 128 runs are reported below. This table corresponds to Table 3 in the main results section, in which only the medians were reported for the sake of conciseness.

| Scenario | | | Total Infections per 100'000 PD | Infections Averted per 100'000 PD | Percent of Infections Averted | Infections Averted per Addtl. 1’000 Doses |
|---|---|---|---|---|---|---|
| *Hospitalization Nudge Prob.* | *Breakthrough Nudge Prob.* | *Miscellaneous Nudge Prob.* | *n* | *n* | *%* | *n* |
| Reference | Reference | Reference | 131.1 (129.9, 132.3) | - | - | . |
| 200% of Reference | Reference | Reference | 125.8 (124.7, 127.0) | 5.3 (4.0, 6.3) | 4.0 (3.1, 4.8) | 391.0 (296.2, 467.2) |
| Reference | 50% of Reference | Reference | 131.2 (130.1, 132.5) | -0.2 (-1.4, 1.0) | -0.1 (-1.1, 0.7) | -232.0 (-2’885.7, 2’959.4) |
| Reference | 0% of Reference | Reference | 131.1 (129.9, 132.7) | 0.0 (-1.6, 1.1) | 0.0 (-1.2, 0.9) | 1’296.0 (-1’249.6, 3’674.6) |
| 200% of Reference | 50% of Reference | Reference | 125.7 (124.8, 127.2) | 5.3 (3.9, 6.3) | 4.1 (2.9, 4.8) | 388.6 (283.9, 459.0) |
| 200% of Reference | 0% of Reference | Reference | 125.8 (124.4, 127.4) | 5.2 (3.7, 6.6) | 4.0 (2.8, 5.0) | 377.9 (265.0, 477.5) |
| Reference | Reference | 50% of Reference | 116.0 (114.5, 119.7) | 15.0 (11.3, 16.6) | 11.4 (8.6, 12.6) | 394.1 (351.8, 420.2) |
| Reference | Reference | 0% of Reference | 100.1 (98.6, 101.6) | 30.9 (29.4, 32.4) | 23.6 (22.4, 24.7) | 391.8 (377.6, 408.2) |
| 200% of Reference | Reference | 50% of Reference | 112.1 (110.1, 119.1) | 18.9 (11.9, 21.0) | 14.4 (9.1, 16.0) | 392.8 (355.3, 419.4) |
| 200% of Reference | Reference | 0% of Reference | 99.8 (98.1, 101.3) | 31.3 (29.8, 33.0) | 23.9 (22.7, 25.2) | 396.2 (378.6, 410.1) |

**Table S5. Overall death rate and deaths averted for select scenarios, with 50% simulation intervals.** For each scenario, the median, 25th percentile, and 75th percentile of each outcome across 128 runs are reported below. This table corresponds to Table 4 in the main results section, in which only the medians were reported for the sake of conciseness.

| Scenario | | | Total Deaths per 100'000 PD | Deaths Averted per 100'000 PD | Percent of Deaths Averted | Deaths Averted per Addtl. 1’000 Doses |
|---|---|---|---|---|---|---|
| *Hospitalization Nudge Prob.* | *Breakthrough Nudge Prob.* | *Miscellaneous Nudge Prob.* | *n* | *n* | *%* | *n* |
| Reference | Reference | Reference | 0.363 (0.349, 0.384) | - | - | - |
| 200% of Reference | Reference | Reference | 0.344 (0.328, 0.362) | 0.018 (0.000, 0.035) | 5.1 (0.1, 9.6) | 1.4 (0.0, 2.7) |
| Reference | 50% of Reference | Reference | 0.364 (0.345, 0.376) | -0.002 (-0.013, 0.018) | -0.4 (-3.6, 5.0) | 9.2 (-24.7, 58.0) |
| Reference | 0% of Reference | Reference | 0.360 (0.343, 0.381) | 0.003 (-0.018, 0.020) | 0.7 (-5.0, 5.5) | 10.3 (-35.9, 57.6) |
| 200% of Reference | 50% of Reference | Reference | 0.346 (0.329, 0.363) | 0.017 (0.000, 0.034) | 4.6 (-0.1, 9.3) | 1.3 (0.1, 2.6) |
| 200% of Reference | 0% of Reference | Reference | 0.342 (0.330, 0.362) | 0.020 (0.000, 0.033) | 5.6 (0.0, 9.1) | 1.4 (0.0, 2.4) |
| Reference | Reference | 50% of Reference | 0.305 (0.288, 0.323) | 0.058 (0.040, 0.075) | 15.9 (10.9, 20.6) | 1.5 (1.1, 2.1) |
| Reference | Reference | 0% of Reference | 0.254 (0.240, 0.266) | 0.108 (0.096, 0.122) | 29.8 (26.5, 33.7) | 1.4 (1.2, 1.5) |
| 200% of Reference | Reference | 50% of Reference | 0.303 (0.287, 0.320) | 0.060 (0.043, 0.076) | 16.5 (11.8, 21.0) | 1.2 (0.9, 1.6) |
| 200% of Reference | Reference | 0% of Reference | 0.250 (0.235, 0.268) | 0.112 (0.094, 0.128) | 31.0 (26.0, 35.2) | 1.4 (1.2, 1.6) |

**Figure S1. Vaccines administered, incidence rate, and death rate by model scenario targeted towards older adults.** For agents aged 65+ years, the hospitalization nudge probability (Hosp. NP) was increased from 100% to 200% of its reference value, the breakthrough nudge probability (Breakthrough NP) was decreased from 100% to 0% of its reference value, and the miscellaneous nudge probabilities (Misc. NP) were decreased from 100% to 0% of their reference values, all in increments of 10%. (For 18- to 64-year-old agents, all probabilities were fixed at their reference values.) For each parameter combination, the median number of vaccine doses administered per run, the median infection rate per 100’000 person-days, and the median disease-related death rate per 100’000 person-days across 128 runs are displayed.

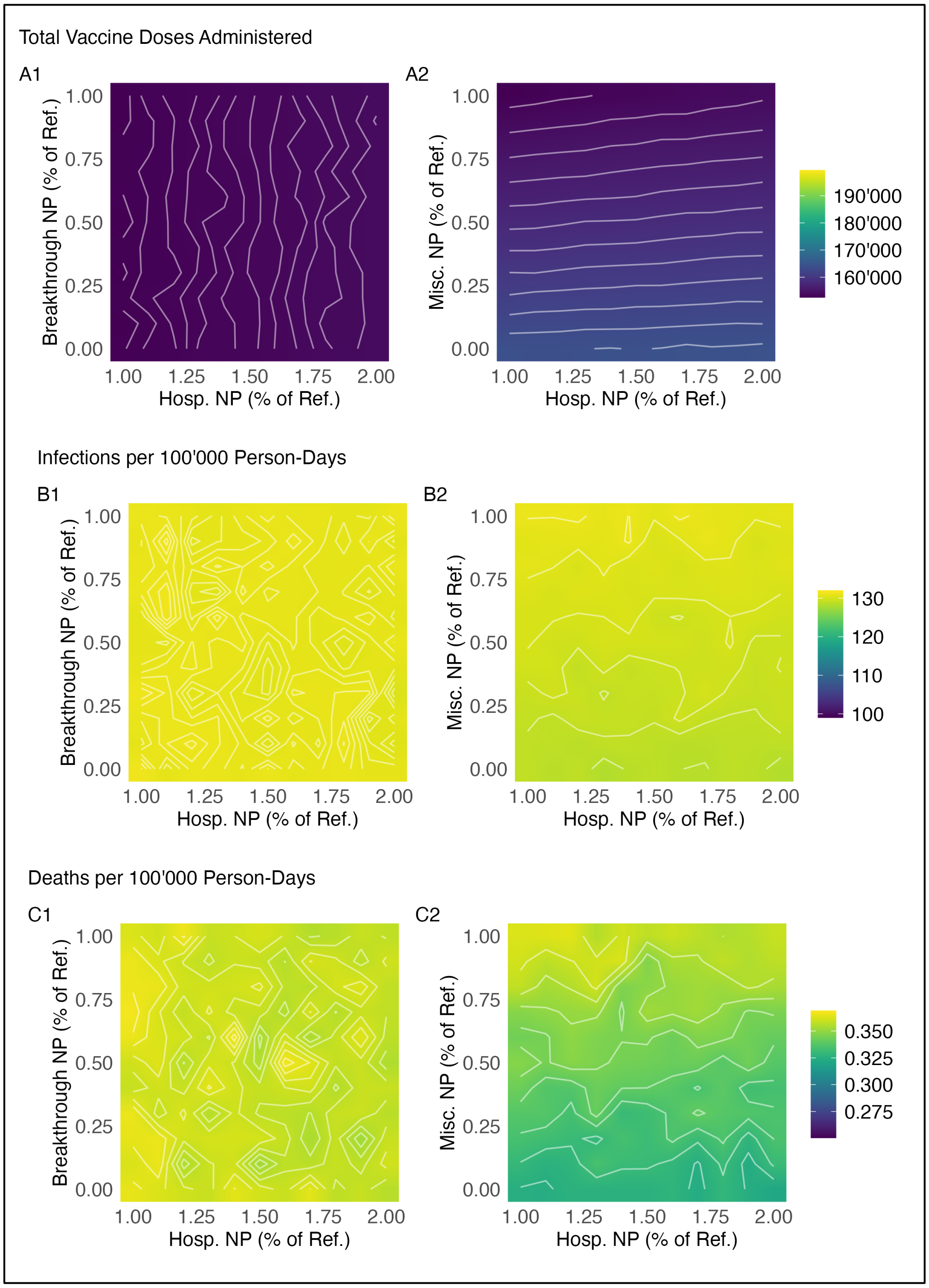

**Figure S2. Vaccines administered, incidence rate, and death rate by model scenario targeted towards younger adults.** For 18- to 64-year-old agents, the hospitalization nudge probability (Hosp. NP) was increased from 100% to 200% of its reference value, the breakthrough nudge probability (Breakthrough NP) was decreased from 100% to 0% of its reference value, and the miscellaneous nudge probabilities (Misc. NP) were decreased from 100% to 0% of their reference values, all in increments of 10%. (For 65+-year-old agents, all probabilities were fixed at their reference values.) For each parameter combination, the median number of vaccine doses administered per run, the median infection rate per 100’000 person-days, and the median disease-related death rate per 100’000 person-days across 128 runs are displayed.

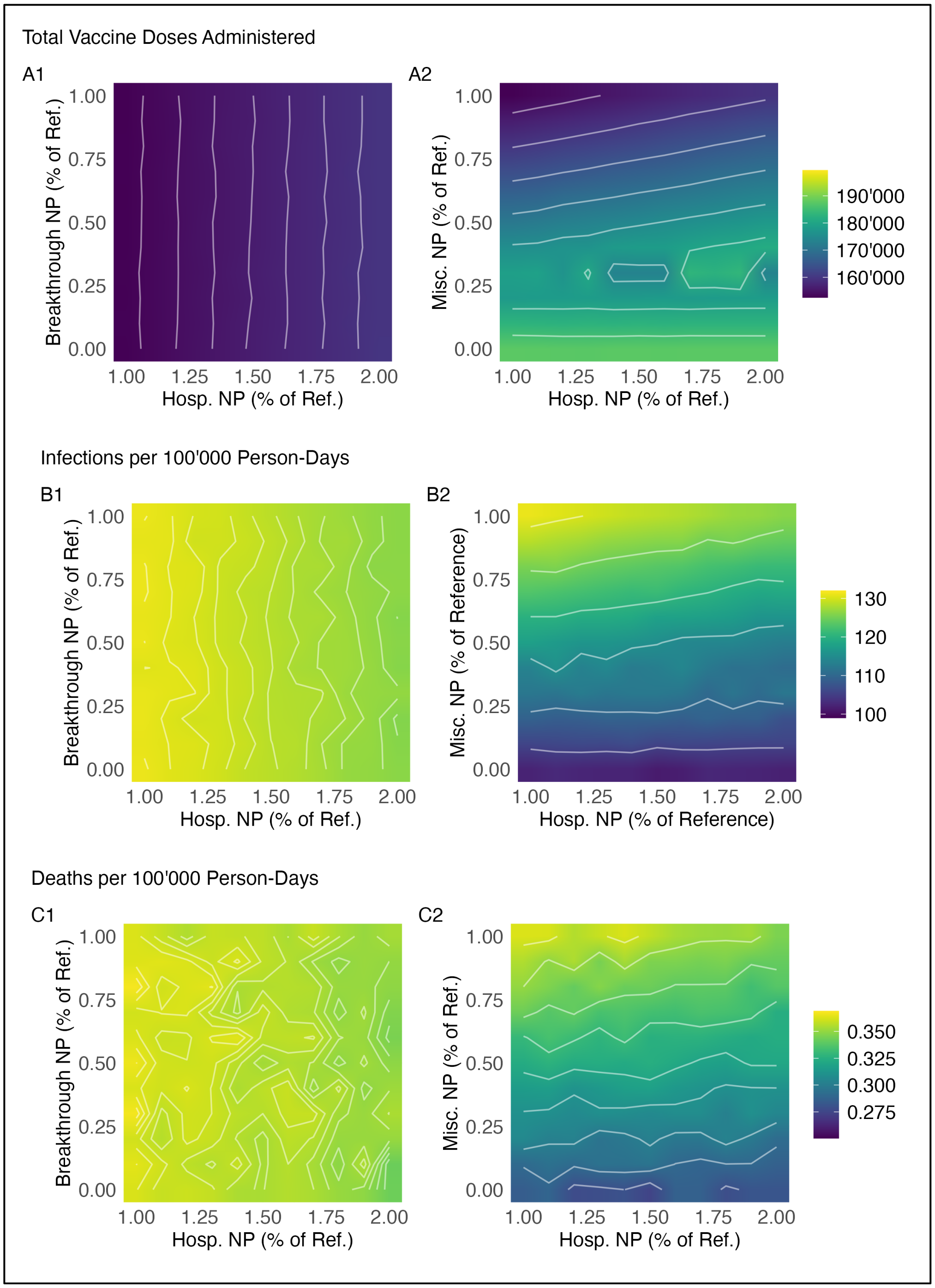

26. Georgia Department of Public Health. *Georgia COVID-19 Dashboard*. Retrieved from https://ga-covid19.ondemand.sas.com/

27. Institute for Health Metrics and Evaluation, University of Washington. (2022, December 16). *IHME: Covid-19 Projections*. Retrieved from https://covid19.healthdata.org/united-states-of-america

28. Centers for Disease Control and Prevention. (2021, May 24). *Covid-19 vaccinations in the United States, jurisdiction*. Retrieved from https://data.cdc.gov/Vaccinations/COVID-19-Vaccinations-in-the-United-States-Jurisdi/unsk-b7fc
